# Tau-vulnerable medial temporal lobe connectivity drives a functional-to-synaptic cascade in Alzheimer’s disease

**DOI:** 10.64898/2026.11.10.26367708

**Authors:** Yijin Zhou, Feng Han, Yihan Ma, Qian Lu, Yihan Fang, Keyan Lou, Dan Wu, Ying Shen, Zhiyong Zhao, the Alzheimer’s Disease Neuroimaging Initiative

## Abstract

Distinct tau-vulnerable and relatively spared subregions exist within the medial temporal lobe (MTL). However, it remains unclear that how functional connectivity (FC), anchored to tau-vulnerable MTL subregions, is related to tau pathology, synaptic dysfunction, and cognitive decline across the Alzheimer’s disease (AD) continuum. We here probe into this with resting-state fMRI data from a discovery cohort (ADNI; N = 386; cognitively normal [CN], mild cognitive impairment [MCI], and AD; 144 with 2-year follow-up) and an independent validation cohort at local hospital (N = 100; CN, subjective cognitive decline [SCD], and MCI; 25 receiving high-frequency repetitive transcranial magnetic stimulation [HF-rTMS]). We first separated the MTL into subregions with high and low neurofibrillary tangle (NFT) burden based on histological atlases, and calculated the seed-based whole-brain FC for each subregion. Second, group differences were assessed using ANCOVA, and associations with amyloid-β (Aβ)/tau pathology, synaptic biomarkers, and cognitive performance were examined. Third, serial mediation, cross-lagged, and longitudinal regression analyses were performed to investigate the proposed pathological sequence, with additional evidence from HF-rTMS intervention and autopsy-confirmed Braak staging. Our results showed that, with the development of AD, FC reductions were focused on the high-NFT MTL network, particularly involving the inferior temporal gyrus and precuneus. Baseline tau burden in high-NFT MTL regions was able to predict subsequent FC alterations. Interestingly, FC between the high-NFT MTL and global brain showed region-specific associations with synaptic biomarkers, including negative associations in the cerebellum and positive associations in the precentral gyrus. We also observed a divergent association between the FC and cognition or pathology at different AD stages, shifting from positive in the early stages to negative in advanced AD. Serial mediation analyses supported a pathway linking tau burden to cognitive impairment through altered high-NFT MTL connectivity in the precuneus and occipital regions and subsequent synaptic dysfunction. Cross-lagged and longitudinal analyses further confirmed the proposed temporal sequence. Furthermore, HF-rTMS-related cognitive improvements were partially associated with enhanced synaptic plasticity through changes in precuneus–high-NFT MTL connectivity. Collectively, these findings support a spatiotemporal framework linking the dysfunction of tau-vulnerable MTL network to synaptic impairment and cognitive decline, highlighting high-NFT MTL-based FC as a potential imaging marker tracking AD progression.

## 1. INTRODUCTION

Alzheimer’s disease (AD) encompasses a progressive clinical continuum, ranging from subjective cognitive decline (SCD) and mild cognitive impairment (MCI) to AD dementia [1]. As the preclinical and prodromal phases of AD, SCD and MCI provide critical windows for early detection and therapeutic intervention [2–4]. The neuropathological progression of AD is characterized by the accumulation of amyloid-β (Aβ) plaques and tau-associated neurofibrillary tangles (NFTs) [5]. Compared with Aβ pathology, the spatiotemporal progression of tau pathology is more closely associated with cognitive deterioration and follows a characteristic anatomical distribution [6, 7]. According to Braak staging, the medial temporal lobe (MTL) is among the earliest regions affected by tau pathology, although its subregions exhibit heterogeneous vulnerability to tau accumulation [8, 9]. Postmortem three-dimensional histological atlases have further revealed that specific MTL subregions accumulate disproportionately high NFT burdens during the earliest disease stages, whereas adjacent regions remain relatively spared until more advanced stages [10, 11]. These findings challenge the conventional view of the MTL as a homogeneous structure and suggest that analyses treating the MTL as a single region may lack the sensitivity to detect early pathological alterations and their effects on functional network integrity.

Previous studies have demonstrated that alterations in functional connectivity (FC), particularly within MTL networks, are sensitive to early neurodegenerative changes [9, 12, 13]. Notably, tau pathology preferentially accumulates and spreads across functionally connected brain regions, suggesting that FC may contribute to the spatial progression of tau pathology along the AD continuum [14, 15]. Therefore, FC alterations may provide a valuable framework for characterizing progressive network dysfunction and elucidating the relationship between regional pathological accumulation and cognitive decline [5, 7, 16].

Increasing evidence implicates synaptic dysfunction in AD progression, linking pathological protein accumulation to neuronal loss and structural atrophy [17–19]. Growth-associated protein 43 (GAP-43), a presynaptic membrane protein abundantly expressed in the hippocampus and entorhinal cortex, has emerged as a sensitive molecular marker of presynaptic remodeling and plasticity [20, 21]. Elevated cerebrospinal fluid (CSF) GAP-43 levels have been observed in individuals with Aβ pathology, suggesting early presynaptic alterations associated with amyloid accumulation [22, 23]. Beyond molecular biomarkers, long-term potentiation (LTP)-like plasticity provides a neurophysiological measure of the brain’s capacity for activity-dependent synaptic strengthening, a fundamental mechanism underlying learning and memory [24]. Impaired LTP-like plasticity reflects disrupted synaptic function and has been associated with memory and executive dysfunction across different stages of AD [25, 26]. Although these findings highlight the critical role of synaptic dysfunction in AD pathogenesis and progression [27], the temporal relationships and potential directional pathways linking resting-state FC alterations, synaptic dysfunction, and subsequent cognitive decline remain poorly understood.

To address these knowledge gaps, the present study integrated multimodal neuroimaging, as well as molecular and neurophysiological biomarkers across two independent cohorts (Fig. 1). Specifically, we aimed to (1) characterize FC alterations associated with pathologically defined high-NFT and low-NFT MTL subregions across the AD continuum; (2) investigate the relationships and potential sequential pathways linking Aβ and tau pathology, MTL-based network dysfunction, synaptic impairment, and cognitive decline; and (3) validate these in vivo functional signatures against autopsy-confirmed Braak neuropathological staging. We hypothesized that high-NFT MTL-based FC would be associated with neuropathological burden, and that FC alterations and synaptic dysfunction would jointly mediate the relationship between AD pathology and cognitive decline. In addition, although high-frequency repetitive transcranial magnetic stimulation (HF-rTMS) has shown potential to improve cognitive performance and to modulate FC and synaptic plasticity in early AD [28–31], the neurobiological mechanisms underlying these effects remain poorly understood. Therefore, we further examined an interventional subset to investigate whether HF-rTMS-related cognitive improvements were associated with changes in MTL-based FC and synaptic plasticity, and whether these changes statistically mediated improvements in cognitive performance.

**Fig. 1.**
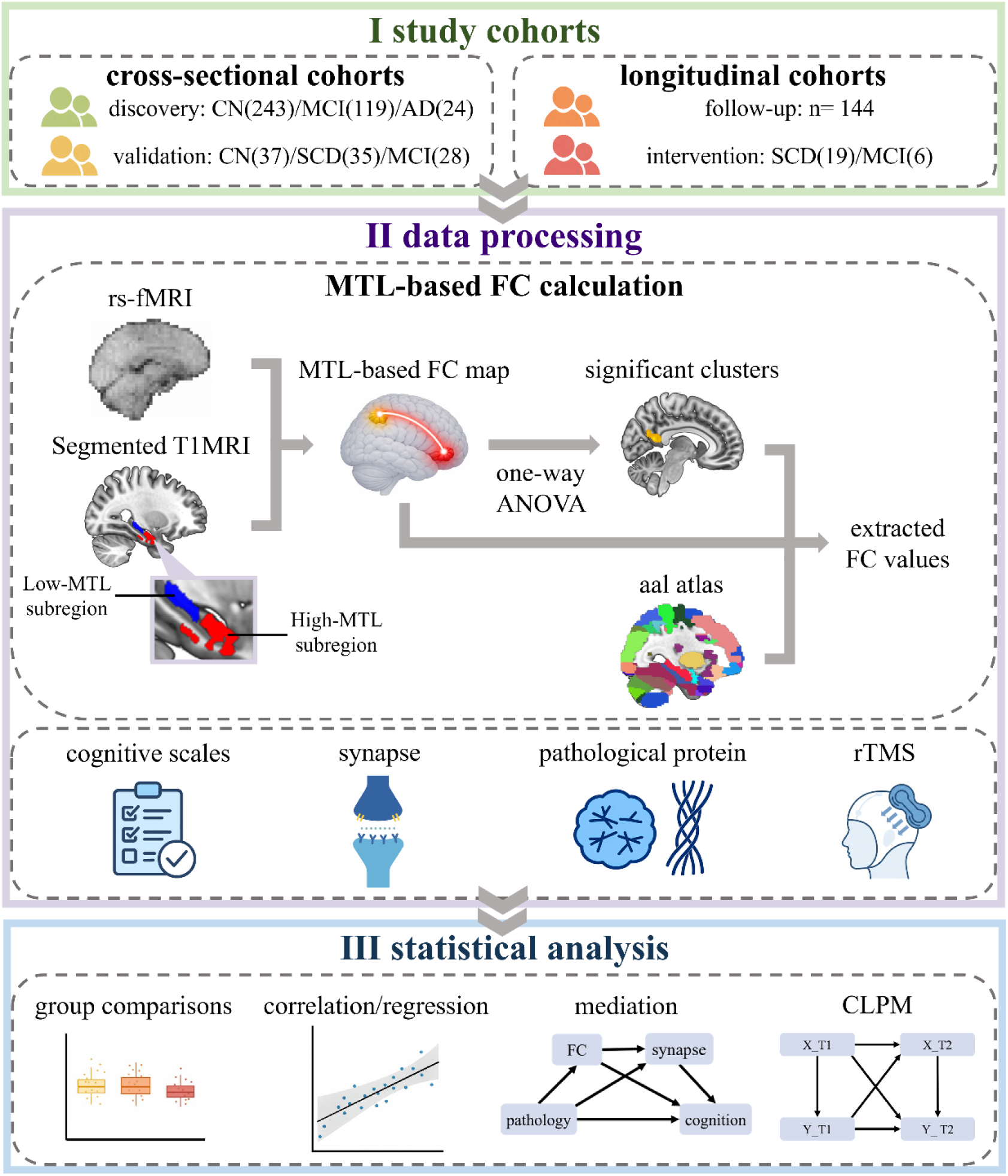
Overview of the study design and analytical workflow. Cross-sectional and longitudinal cohorts were used to investigate functional connectivity (FC) patterns associated with medial temporal lobe (MTL) regions characterized by high or low neurofibrillary tangle (NFT) burden. Multimodal assessments integrated cognitive performance, synaptic biomarkers (GAP-43 and LTP-like plasticity), measures of AD pathology, and neuropathological findings. The analytical framework included cross-sectional modeling of a hypothesized pathological cascade, longitudinal cross-lagged panel modeling (CLPM) to examine temporal relationships among key variables, and neuropathological validation using postmortem Braak staging. CN: cognitively normal, MCI: mild cognitive impairment, AD: Alzheimer’s disease, SCD: subjective cognitive decline, rs-fMRI: resting-state functional magnetic resonance imaging, T1MRI: T1-weighted magnetic resonance imaging, MTL: medial temporal lobe, FC: functional connectivity, NFT, neurofibrillary tangle; GAP-43, growth-associated protein 43; LTP, long-term potentiation; ANOVA: analysis of variance, AAL: Automated Anatomical Labeling, rTMS: repetitive transcranial magnetic stimulation, CLPM: cross-lagged panel model.

## 2. METHODS

### 2.1 Participants

Data for the discovery cohort were obtained from the Alzheimer’s Disease Neuroimaging Initiative (ADNI) database. Participants met the standardized ADNI inclusion criteria, including an age of 55–90 years, a Hachinski Ischemic Scale score ≤ 4, a Geriatric Depression Scale score (GDS) < 6, and no medical contraindications to magnetic resonance imaging (MRI). From an initial sample of 421 participants, 35 were excluded following MRI quality control (i.e. poor spatial normalization, incomplete brain coverage, or excessive head motion), resulting in a final discovery cohort of 386 participants with both T1-weighted structural MRI and resting-state functional MRI (rs-fMRI) data. These included 243 cognitively normal (CN) participants, 119 individuals with MCI, and 24 patients with AD, classified according to ADNI diagnostic criteria. CSF GAP-43 measurements were available for a subset of 157 participants. Longitudinal analyses included 144 participants with both T1-weighted MRI and rs-fMRI data available at two or more time points. The ADNI study was conducted in accordance with the Declaration of Helsinki, and its protocol was approved by the Institutional Review Boards (IRBs; http://adni.loni.usc.edu/wp-content/uploads/2013/09/DOD-ADNI-IRB-Approved-Final-protocol-08072012.pdf) of all participating institutions (ADNI study protocol). Written informed consent was obtained from all participants before enrollment.

The independent external validation cohort was recruited from the Department of Rehabilitation Medicine at Jiangsu Shengze Hospital, affiliated with Nanjing Medical University. The cohort comprised 100 participants, including 37 CN individuals, 35 individuals with SCD, and 28 individuals with MCI. A subset of 25 participants (19 with SCD and 6 with MCI) was included in the intervention study. The study was approved by the local Ethics Committee (No. JSSZYY-LLSC-202020) and conducted in accordance with the Declaration of Helsinki. The study was registered with the Chinese Clinical Trial Registry (No. ChiCTR2100046131). Written informed consent was obtained from all participants. Eligible participants were aged 55–80 years. Exclusion criteria included vascular dementia, drug or alcohol dependence, contraindications to MRI, TMS and severe medical, neurological, or psychiatric conditions, including clinically significant depressive symptoms (GDS score ≥ 6). Participants were classified based on comprehensive neuropsychological assessments and subjective cognitive complaints, following the diagnostic criteria and eligibility requirements described in our previous study [26].

### 2.2 Clinical scale assessments

In the discovery cohort, global cognitive function was assessed using the Mini-Mental State Examination (MMSE) and Montreal Cognitive Assessment (MoCA), while the 13-item Alzheimer’s Disease Assessment Scale–Cognitive Subscale (ADAS13) was used to evaluate cognitive impairment in greater detail. Clinical disease severity was assessed using the Clinical Dementia Rating–Sum of Boxes (CDR-SB) scale, and functional abilities in daily living were evaluated using the FAQ. Language function, particularly confrontation naming, was assessed using the Multilingual Naming Test (MINT). Depressive symptoms were evaluated using the GDS

In the validation cohort, participants underwent a comprehensive neuropsychological battery assessing multiple cognitive domains, as described in our previous study [26]. Detailed descriptions of the assessments are provided in the Supplementary Materials.

### 2.3 MRI acquisition

In the discovery cohort, rs-fMRI data obtained from the ADNI database were acquired using 3T scanners manufactured by GE, Philips, or Siemens, whereas T1-weighted structural images were acquired using 1.5T or 3.0T scanners from the same manufacturers. Rs-fMRI data were acquired using echo-planar imaging (EPI) sequences. Owing to the multisite nature of ADNI, acquisition parameters varied across participants. Repetition times (TRs) were approximately 1,500, or 3,000 ms, and the number of volumes was ∼200. The number of slices was 48, 51, or 64, with voxel dimensions ranging from 2.5 × 2.5 × 2.5 mm³ to 3.44 × 3.44 × 3.4 mm³. T1-weighted structural images were acquired using three-dimensional magnetization-prepared rapid gradient-echo (MPRAGE) or inversion recovery–spoiled gradient-recalled acquisition in steady state (IR-SPGR) sequences. Acquisition parameters were as follows: TR = 6.4–2534.7 ms, echo time (TE) = 2.9–3.2 ms, inversion time (TI) = 0, 400, or 900 ms, flip angle = 9° or 11°, and slice thickness = 1.0 or 1.2 mm. Detailed MRI acquisition protocols and related documentation are available on the ADNI website (https://adni.loni.usc.edu).

In the validation cohort, neuroimaging data were acquired using a 3.0T GE Discovery MR750w (SIGNA) MRI scanner equipped with a 24-channel head coil. High-resolution T1-weighted structural images were acquired using a three-dimensional spoiled gradient-recalled echo (3D T1-SPGR) sequence (voxel size = 1 × 1 × 1 mm³; TR/TE = 8.7/3.2 ms). Rs-fMRI data were acquired using a gradient-echo EPI sequence with 41 interleaved slices (voxel size = 3 × 3 × 3.5 mm³; TR/TE = 2,500/30 ms). Additional details of the MRI acquisition protocol have been reported in our previous study [26].

### 2.4 MRI processing

Identical MRI preprocessing and structural segmentation pipelines were applied to the discovery and validation cohorts. Rs-fMRI data were preprocessed using the Data Processing Assistant for Resting-State fMRI, Advanced Edition (DPARSFA, version 5.5), implemented in MATLAB R2024b [32]. The preprocessing pipeline comprised the following steps: (1) removal of the first 10 volumes; (2) slice-timing correction; (3) realignment to correct for head motion, with participants exhibiting excessive head motion (translation > 3 mm or rotation > 3°) excluded from subsequent analyses; (4) coregistration of structural and functional images; (5) nuisance regression, including the Friston 24-parameter head motion model, white matter and CSF signals, and removal of linear trends. Global signal regression was not performed because of concerns regarding its potential to introduce spurious negative correlations [33]; (6) spatial normalization to Montreal Neurological Institute (MNI) space using EPI templates, followed by resampling to an isotropic voxel size of 3 × 3 × 3 mm³ [34]; (7) temporal band-pass filtering (0.01–0.1 Hz) to attenuate low-frequency drift and high-frequency physiological noise; and (8) spatial smoothing with a Gaussian kernel of 6-mm full width at half maximum (FWHM).

T1-weighted structural images were processed to delineate MTL subregions using the Automatic Segmentation of Hippocampal Subfields pipeline with the Penn Memory Center T1 atlas (ASHS-PMC-T1) [35, 36]. To account for regional differences in vulnerability to tau pathology, the segmented MTL subregions were classified into high-and low-NFT regions based on a validated three-dimensional histopathological atlas of NFT density. This atlas was derived from postmortem data from 15 individuals and thresholded to retain regions in which at least 20% of individuals exhibited mild or greater NFT burden [9]. The high-NFT region of interest (ROI) encompassed the entorhinal cortex (ERC) and Brodmann area 35 (BA35), representing regions particularly vulnerable to early tau accumulation. The low-NFT ROI was defined by excluding the high-NFT regions from the total MTL segmentation and primarily comprised the anterior and posterior hippocampus and parahippocampal cortex (PHC), which are relatively spared during the earliest disease stages [11]. Finally, spatial normalization was performed using Advanced Normalization Tools (ANTs), and the resulting ROIs were resampled to match the spatial resolution and alignment of the normalized functional images [37].

### 2.5 MTL-based functional connectivity calculation

In both the discovery and validation cohorts, seed-based FC analyses were performed using the segmented high-NFT and low-NFT ROIs as seeds. For each participant, the mean blood oxygen level-dependent (BOLD) time series was extracted from each seed ROI. Voxel-wise Pearson correlation coefficients were then calculated between each seed time series and the time series of all voxels within the whole-brain gray matter mask. The resulting correlation maps were converted to Fisher’s z-score maps to improve the normality of the correlation coefficients for subsequent group-level statistical analyses.

### 2.6 Synaptic biomarkers

In the discovery cohort, CSF GAP-43 concentrations (pg/mL) were quantified using an enzyme-linked immunosorbent assay (ELISA). The assay employed a mouse monoclonal antibody (NM4) for coating and a polyclonal antibody (ABB-135) for detection, specifically recognizing the C-terminal of the GAP-43 protein.

In the validation cohort, synaptic function was assessed using LTP-like plasticity induced by intermittent theta-burst stimulation (iTBS) applied over the left motor cortex, following previously established protocols [26, 38, 39]. Motor-evoked potentials (MEPs) were recorded from the right abductor pollicis brevis (APB) muscle at baseline and at 5, 10, and 30 min after stimulation using a D-MT500 magnetic stimulator. LTP-like plasticity at each post-stimulation time point (LTP-5min, LTP-10min, and LTP-30min) was quantified as the percentage change in MEP amplitude relative to baseline.

### 2.7 Pathological measures

In the discovery cohort, in vivo Aβ and tau pathology were assessed using positron emission tomography (PET). Regional standardized uptake value ratio (SUVR) data were obtained from the ADNI database. Aβ PET imaging was performed using [¹⁸F]florbetapir, [¹⁸F]florbetaben, or [¹⁸F]NAV4694, whereas tau PET imaging was performed using [¹⁸F]flortaucipir (AV-1451). Regional SUVRs corresponding to Braak stages 1, 3/4, and 5/6 were calculated for both Aβ and tau using FreeSurfer-defined ROI. The Braak stage 2 composite was excluded from all analyses because of potential contamination from off-target [¹⁸F]flortaucipir binding in the adjacent choroid plexus. As the cerebellum served as the reference region for calculating both Aβ and tau SUVRs, it was excluded from pathology-related analyses. Among the 386 participants, in-vivo Aβ PET and tau PET data were available for 320 and 237 individuals, respectively.

In the validation cohort, serum Aβ1–42 and phosphorylated tau at threonine 181 (p-tau181) concentrations were quantified using a double-antibody sandwich ELISA. Serum samples were added to microtiter plates pre-coated with specific monoclonal capture antibodies. After washing to remove unbound components, polyclonal detection antibodies were added, followed by horseradish peroxidase (HRP)-conjugated secondary antibodies to form antibody–antigen–antibody–HRP complexes. Following termination of the substrate reaction, optical density (OD) was measured at 450 nm. Analyte concentrations were calculated using SoftMax Pro software based on quadratic standard curves relating concentration to absorbance. For subsequent analyses, concentrations were expressed in units of 1,000 pg/L. Among the 100 participants, serum pathology measurements were available for 80 individuals.

To complement and validate the in vivo neuroimaging findings, an independent subset of 19 participants with available postmortem neuropathological data was identified from the ADNI database. Neuropathological Braak stages were extracted based on the postmortem distribution and severity of NFT pathology. Neuropathological assessments were performed in accordance with the National Institute on Aging–Alzheimer’s Association (NIA-AA) guidelines, with findings documented using the National Alzheimer’s Coordinating Center (NACC) Neuropathology Data Forms.

### 2.8 Repetitive TMS intervention

A subset of 25 participants (19 with SCD and 6 with MCI) from the validation cohort underwent a 4-week HF-rTMS intervention. Stimulation targeted the left dorsolateral prefrontal cortex (L-DLPFC), identified using the F3 position of the international 10–20 electroencephalography (EEG) system. Magnetic pulses were delivered using a D-MT500 stimulator equipped with a figure-of-eight coil (Neurosoft Ltd., Russia). The HF-rTMS protocol consisted of stimulation at 10 Hz and an intensity of 90% of the resting motor threshold (RMT). Each session comprised 40 trains of 5 s each, separated by 10-s intertrain intervals, delivering a total of 2,000 pulses. The intervention was administered five days per week for four consecutive weeks, comprising 20 treatment sessions. Participants were monitored for potential adverse events throughout the intervention period.

The intervention was conducted at Jiangsu Shengze Hospital between May and December 2021, with approval from the hospital’s IRB. All procedures were performed in accordance with the Declaration of Helsinki.

### 2.9 Statistical analyses

Statistical analyses were performed using R (version 4.5.1). Age, sex, and years of education were included as covariates in group comparisons, regression analyses, mediation analyses, and cross-lagged panel models (CLPM). The interval between MRI acquisition and CSF sampling was additionally included as a covariate in GAP-43-related analyses, whereas head motion during rs-fMRI acquisition was controlled for in FC analyses. Raw CSF GAP-43 concentrations were log10-transformed to reduce skewness and improve their suitability for statistical analyses [40, 41].

Cross-sectional FC analyses were conducted at both the voxel and ROI levels in the discovery and validation cohorts. Group differences in FC were assessed using one-way analysis of covariance (ANCOVA), followed by post hoc pairwise comparisons to identify differences between specific diagnostic groups. For ROI-level analyses, mean FC values were extracted using the Automated Anatomical Labeling (AAL) atlas. Site-related effects were harmonized using the ComBat algorithm.

Subsequently, multiple linear regression analyses were performed to examine associations between neurobiological features and clinical or pathological measures. The general regression model was specified as follows:

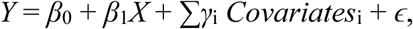

For analyses of clinical associations, Y represents cognitive performance, and X denotes neurobiological features, including FC measures or synaptic biomarkers. For pathology-related analyses, Y represents neurobiological features, whereas X denotes pathological burden, as measured by Aβ or tau biomarkers. Covariates represent the previously specified control variables. Unstandardized regression coefficients (*β*) and their corresponding P values were reported.

Finally, simple and serial mediation analyses were performed using structural equation modeling (SEM). Simple mediation models examined the indirect associations between synaptic biomarkers and cognitive outcomes (MMSE and MoCA) through FC, as well as between FC and cognitive outcomes through synaptic biomarkers. Serial mediation models evaluated pathways linking Aβ and tau pathology to cognitive outcomes through FC and synaptic biomarkers, considering both possible orders of these mediators. Indirect effects were estimated using 5,000 bootstrap resamples and were considered statistically significant when their 95% bootstrap confidence intervals (CIs) did not include zero.

For the longitudinal dataset, two-wave CLPMs were constructed within a SEM framework to examine the temporal and directional associations among pathological burden, synaptic biomarkers, FC, and cognitive performance. The models included autoregressive paths to account for the temporal stability of each variable and cross-lagged paths to assess whether baseline levels of one variable predicted subsequent changes in another, after accounting for its baseline level. Model parameters were estimated using maximum likelihood estimation, with 5,000 bootstrap resamples used to assess statistical uncertainty. Standardized regression coefficients (β) and corresponding P values were reported.

For the intervention analyses, statistical assessments were performed separately for the overall intervention cohort and the SCD subgroup. The MCI subgroup was not analyzed separately because of its limited sample size. Paired t-tests were used to assess pre- to post-intervention changes in clinical scale scores, LTP-like plasticity, and FC measures in regions identified as showing significant alterations in the cross-sectional analyses. To quantify intervention-related changes, difference scores (post-intervention minus baseline) were calculated and subsequently incorporated into mediation models to examine whether changes in synaptic function mediated the association between FC changes and cognitive improvements. The statistical significance of indirect effects was assessed using 5,000 bootstrap resamples, with effects considered significant when the corresponding 95% CIs excluded zero.

To validate the in vivo neuroimaging findings against postmortem neuropathological evidence, additional analyses were performed using autopsy-derived neuropathological data. Linear correlation analyses and the Sasabuchi–Lind–Mehlum (SLM) test were conducted to examine the relationships between FC in the identified significant clusters and neuropathological Braak stages, including potential nonlinear associations. Participants with available autopsy data were further stratified into three groups according to Braak stage: stages 1-2, stages 3-4, and stages 5-6. Group differences in FC within the significant clusters were assessed using ANCOVA, adjusting for the previously specified demographic and clinical covariates.

To account for multiple comparisons, Gaussian random field (GRF) correction was applied to voxel-wise FC analyses, using a voxel-level threshold of *p* < 0.01 and a cluster-level threshold of *p* < 0.05 (two-tailed) to identify statistically significant clusters. False discovery rate (FDR) correction (*q < 0.05*) was applied where appropriate. For multiple linear regression analyses across clinical stages, FDR correction was performed separately within each diagnostic group. For ROI-level FC analyses based on the AAL atlas, FDR correction was applied across all 116 regions for each statistical model, including cross-sectional correlation analyses and longitudinal CLPMs.

## 3. RESULTS

### 3.1 Demographic and clinical characteristics

As shown Table 1, the discovery cohort exhibited significant group differences in age (χ² = 13.37, *p* = 0.001) and sex distribution (χ² = 8.93, *p* = 0.012), with the AD group generally being older than the other groups and the CN group having a higher proportion of female individuals. In contrast, years of education did not differ significantly among the three groups (*p* = 0.587). Global cognitive performance, as assessed by the MMSE, progressively declined across the clinical spectrum, whereas disease severity, as measured by the CDR-SB, increased significantly (both *p* < 0.001).

**Table 1.** Demographic and clinical characteristics of participants across CN, MCI, and AD groups in the discovery cohort.

|  | CN (N=243) | MCI (N=119) | AD (N=24) | *χ*<sup>2</sup> | *p* |
|---|---|---|---|---|---|
| Sex(F/M) | 148/95 | 53/66 | 12/12 | 8.93 | 0.012 |
| Age | 73.00 (10.00) | 73.00 (10.00) | 79.00 (7.25) | 13.37 | 0.001 |
| Education | 16.00 (3.50) | 16.00 (4.00) | 16.00 (3.25) | 1.06 | 0.587 |
| MMSE (N=385) | 29.00 (2.00) | 28.00 (3.00) | 22.00 (3.25) | 90.39 | <0.001 |
| CDR-SB (N=385) | 0.00 (0.00) | 1.50 (1.50) | 4.75 (4.00) | 284.26 | <0.001 |

Continuous variables are presented as medians (interquartile ranges [IQRs]) and were compared across groups using the Kruskal–Wallis test. Categorical variables (sex) are presented as counts and were compared using Pearson’s chi-square test. The χ² statistic represents the Kruskal–Wallis test statistic for continuous variables and Pearson’s chi-square statistic for categorical variables. CN, cognitively normal; MCI, mild cognitive impairment; AD, Alzheimer’s disease; F/M, female/male; MMSE, Mini-Mental State Examination; CDR-SB: Clinical Dementia Rating–Sum of Boxes; MoCA, Montreal Cognitive Assessment.

In the validation cohort (Supplementary Table 1), no significant group differences were observed in sex distribution (χ² = 0.98, *p* = 0.612) or educational level (χ² = 0.13, *p* = 0.938). However, age differed significantly among the three groups (χ² = 5.88, *p* = 0.001), with the MCI group exhibiting the highest median age. Global cognitive performance, as assessed by the MMSE and MoCA, showed a significant progressive decline across the clinical spectrum (both *p* < 0.001).

### 3.2 Group difference of FC and its association with synaptic biomarkers

Voxel-wise one-way ANCOVA revealed significant reductions in FC between the high-NFT MTL region and brain regions primarily involving the inferior temporal gyrus (ITG) and precuneus after FDR correction (*p* < 0.05). Post hoc analyses demonstrated a progressive decline in high-NFT MTL-based FC from CN participants to those with MCI and AD (Fig. 2a). Consistent with the discovery cohort, the validation cohort showed progressive reductions in FC between the high-NFT MTL region and the middle frontal gyrus (MFG) and precuneus across the CN, SCD, and MCI groups (Fig. 2b). No significant group differences in low-NFT MTL-based FC were detected in either cohort. ROI-level analyses further revealed a significant group difference in FC between the high-NFT MTL region and the left ITG (*p* = 0.001).

**Fig. 2.**
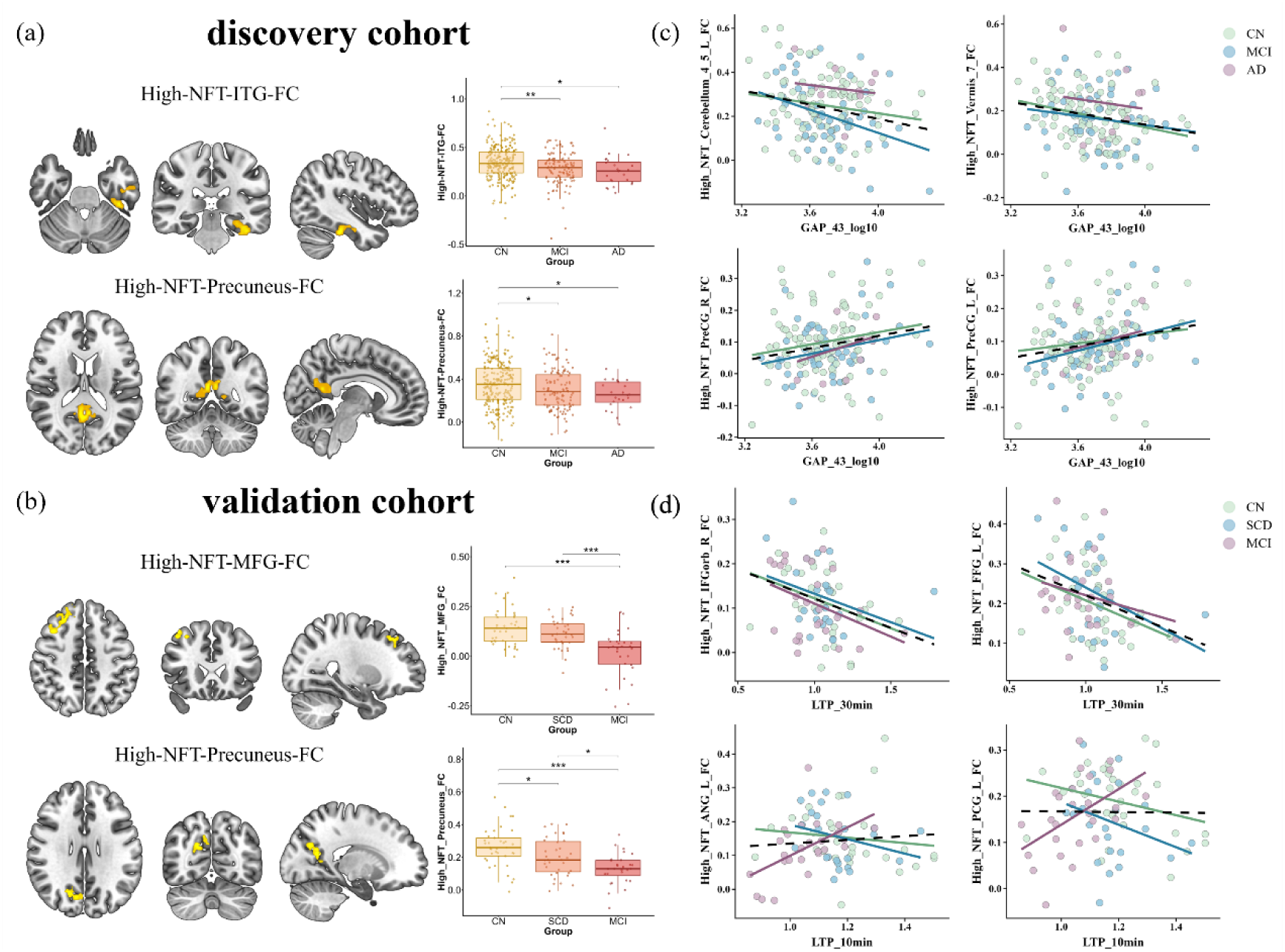
Group differences in high-NFT MTL-based functional connectivity and its associations with synaptic biomarkers. (a) Discovery cohort: Significant reductions in functional connectivity (FC) between the high-NFT medial temporal lobe (MTL) region and the inferior temporal gyrus (ITG) and precuneus across the CN, MCI, and AD groups. (b) Validation cohort: Corresponding reductions in FC between the high-NFT MTL region and the middle frontal gyrus (MFG) and precuneus across the CN, SCD, and MCI groups. (c) Associations between log10-transformed cerebrospinal fluid (CSF) GAP-43 levels and high-NFT MTL-based FC in the discovery cohort. (d) Associations between long-term potentiation (LTP)-like plasticity measured at 30 and 10 min after stimulation (LTP-30min and LTP-10min) and high-NFT MTL-based FC in the validation cohort. In the scatter plots, black dashed lines represent regression fits for the overall sample, whereas colored lines represent regression fits for individual diagnostic groups. All analyses were adjusted for age, sex, and years of education. * *p* < 0.05, ** *p* < 0.01, *** *p* < 0.001. CN, cognitively normal; SCD, subjective cognitive decline; MCI, mild cognitive impairment; AD, Alzheimer’s disease; ITG, inferior temporal gyrus; MFG, middle frontal gyrus; PreCG, precentral gyrus; IFGorb, orbital part of the inferior frontal gyrus; FFG, fusiform gyrus; ANG, angular gyrus; PCG, posterior cingulate gyrus; Cerebellum_4_5, cerebellar lobules IV–V; Vermis_7, cerebellar vermis lobule VII; L, left; R, right.

Across all participants in the discovery cohort, log10-transformed CSF GAP-43 levels showed nominally significant correlations with high-NFT MTL-based FC in several brain regions. Negative correlations were observed in the left cerebellar lobules IV–V (*r* = −0.230, *p* = 0.004) and vermis VI (*r* = −0.201, *p* = 0.012). In contrast, positive correlations were identified in the right (*r* = 0.200, *p* = 0.012) and left precentral gyri (*r* = 0.194, *p* = 0.015). Similar correlation patterns were observed in analyses stratified by diagnostic group (Fig. 2c). In the validation cohort, late-phase LTP-like plasticity, measured at 30 min after stimulation (LTP-30min), showed the strongest negative correlations with high-NFT MTL-based FC. The most pronounced associations were observed in the right orbital part of the inferior frontal gyrus (*r* = −0.327, *p* = 0.003) and left fusiform gyrus (*r* = −0.326, *p* = 0.003). Within the MCI subgroup, early-phase LTP-like plasticity measured at 10 min (LTP-10min) showed nominally significant positive correlations with high-NFT MTL-based FC in the left angular gyrus (*r* = 0.531, *p* = 0.005) and left posterior cingulate region (r = 0.485, *p* = 0.012) (Fig. 2d). However, none of these associations survived FDR correction.

### 3.3 Relationships between FC and clinical and pathological measures

Regression analyses examining FC as the independent variable and clinical scores as the dependent variables revealed stage-dependent differences in the direction of associations. For most cognitive measures in which higher scores indicate better performance, regression coefficients for FC between the high-NFT MTL region and the precuneus were positive in the overall sample and CN subgroup but negative in the AD subgroup. Conversely, for measures in which higher scores indicate greater impairment, the associations shifted from negative to positive across the CN, MCI, and AD groups. Specifically, across the entire sample, FC between the high-NFT MTL region and the ITG was negatively associated with ADAS13 scores (*β* = −7.04, FDR-corrected *p* = 0.029) and positively associated with MMSE scores (*β* = 1.79, FDR-corrected *p* = 0.048). In contrast, within the AD subgroup, greater high-NFT MTL–ITG FC was significantly associated with poorer daily functioning and more severe depressive symptoms, as reflected by higher FAQ scores (*β* = 27.60, FDR-corrected *p* = 0.032) and GDS scores (*β* = 7.72, FDR-corrected *p* = 0.026), respectively (Fig. 3a, c).

**Fig. 3.**
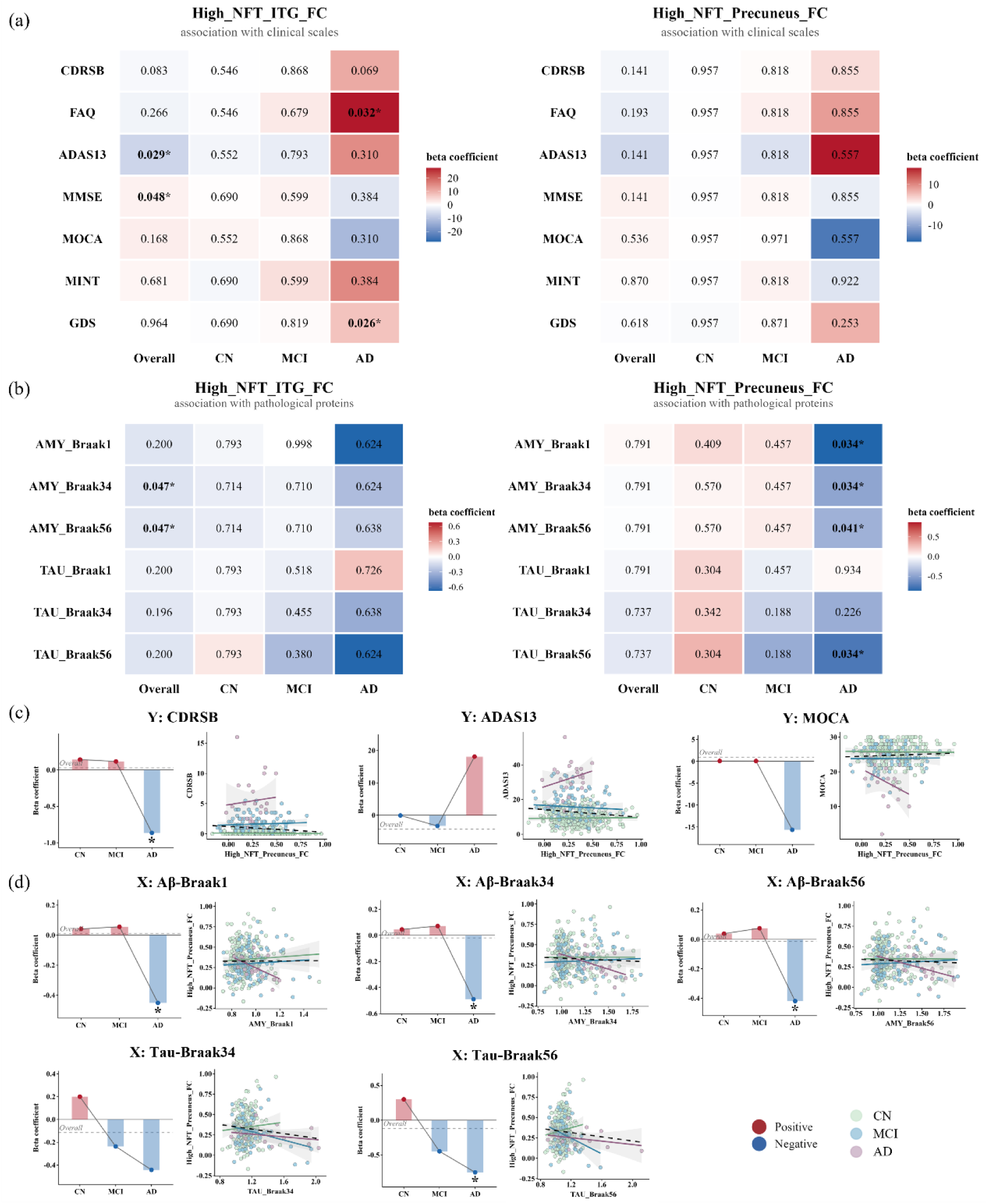
Stage-dependent associations of high-NFT functional connectivity with clinical performance and pathological burden. (a) Heatmaps illustrating associations between high-NFT medial temporal lobe (MTL)-based functional connectivity (FC) with the inferior temporal gyrus (ITG) and precuneus and clinical scale scores across diagnostic groups. Cell values indicate *p* values, and colors represent standardized regression coefficients (*β*). (b) Heatmaps illustrating associations between high-NFT MTL-based FC (ITG and precuneus) and amyloid/tau pathological burden across Braak-stage-defined regions. (c) Changes in regression coefficients for the associations between high-NFT MTL–precuneus FC and clinical scale scores across the CN, MCI, and AD groups. (d) Changes in regression coefficients for the associations between pathological burden and high-NFT MTL–precuneus FC across the CN, MCI, and AD groups. Together, these plots illustrate stage-dependent shifts in the direction and magnitude of the associations as disease progresses. Asterisks (*) indicate significant associations (*p* < 0.05). AMY, amyloid; ADAS13, Alzheimer’s Disease Assessment Scale–Cognitive Subscale; MMSE, Mini-Mental State Examination; MoCA, Montreal Cognitive Assessment; FAQ, Functional Activities Questionnaire; GDS, Geriatric Depression Scale; MINT, Multilingual Naming Test.

Regarding associations with pathological burden, higher Aβ burden in Braak 3/4 (*β* = −0.11, FDR-corrected *p* = 0.047) and Braak5/6 (*β* = −0.10, FDR-corrected *p* = 0.047) regions was significantly associated with lower FC between the high-NFT MTL region and ITG across the entire sample. Furthermore, greater Aβ burden in Braak1 (*β* = −0.86, FDR-corrected *p* = 0.034), Braak3/4 (*β* = −0.49, FDR-corrected *p* = 0.034), and Braak5/6 (*β* = −0.42, FDR-corrected *p* = 0.041) regions—as well as higher tau burden in Braak5/6 regions (*β* = −0.75, FDR-corrected *p* = 0.034), was significantly associated with reduced high-NFT MTL–precuneus FC. Notably, the associations between Aβ and tau burden and high-NFT MTL–precuneus FC showed a reversal in direction across disease stages, shifting from CN to AD (Fig. 3b, d).

Both clinical performance and pathological burden exhibited complex associations with GAP-43 levels (Supplementary Fig.1).

In the validation cohort, regression analyses across all participants revealed widespread significant positive associations between high-NFT MTL-based FC involving the MFG and precuneus and cognitive performance. Stage-dependent reversals in the direction of these associations were also observed across multiple cognitive domains (Supplementary Fig. 2a and Fig. 4a). Regarding pathological biomarkers, serum Aβ1–42 levels were positively associated with high-NFT MTL–precuneus FC (*β* = 0.94, FDR-corrected *p* = 0.032) and high-NFT MTL–MFG FC (*β* = 0.83, FDR-corrected *p* = 0.045) FC within the MCI subgroup. In contrast, a significant negative association was observed between Aβ1–42 levels and high-NFT MTL–precuneus FC in the CN group (*β* = −0.88, FDR-corrected *p* = 0.034), indicating a stage-dependent reversal in the direction of these associations (Supplementary Fig. 2b and Fig. 4b). Given the expected inverse relationship between Aβ1–42 levels and cerebral amyloid plaque burden, this pattern may be consistent with the stage-dependent alterations observed in the discovery cohort [42, 43]. Furthermore, early-phase synaptic efficacy (LTP 10 min) demonstrated robust and highly min after stimulation (LTP-10min), showed significant positive associations with cognitive performance across nearly all assessed domains (Supplementary Fig. 3a and Fig. 3b).

### 3.4 Mediation effect of the MTL-based FC to synaptic biomarkers pathway linking pathological protein to cognition

Serial mediation analyses revealed significant indirect associations between tau burden in Braak 3/4 regions and MMSE scores through sequential alterations in high-NFT MTL-based FC and CSF GAP-43 levels. Specifically, significant serial indirect effects were observed for FC involving the left calcarine cortex (95% CI: 0.36 to 5.16), right precuneus (95% CI: 0.27 to 4.34), and left (95% CI: 0.24 to 4.54) and right superior occipital gyri (SOG; 95% CI: 0.26 to 4.26) (Fig. 4a).

**Fig. 4.**
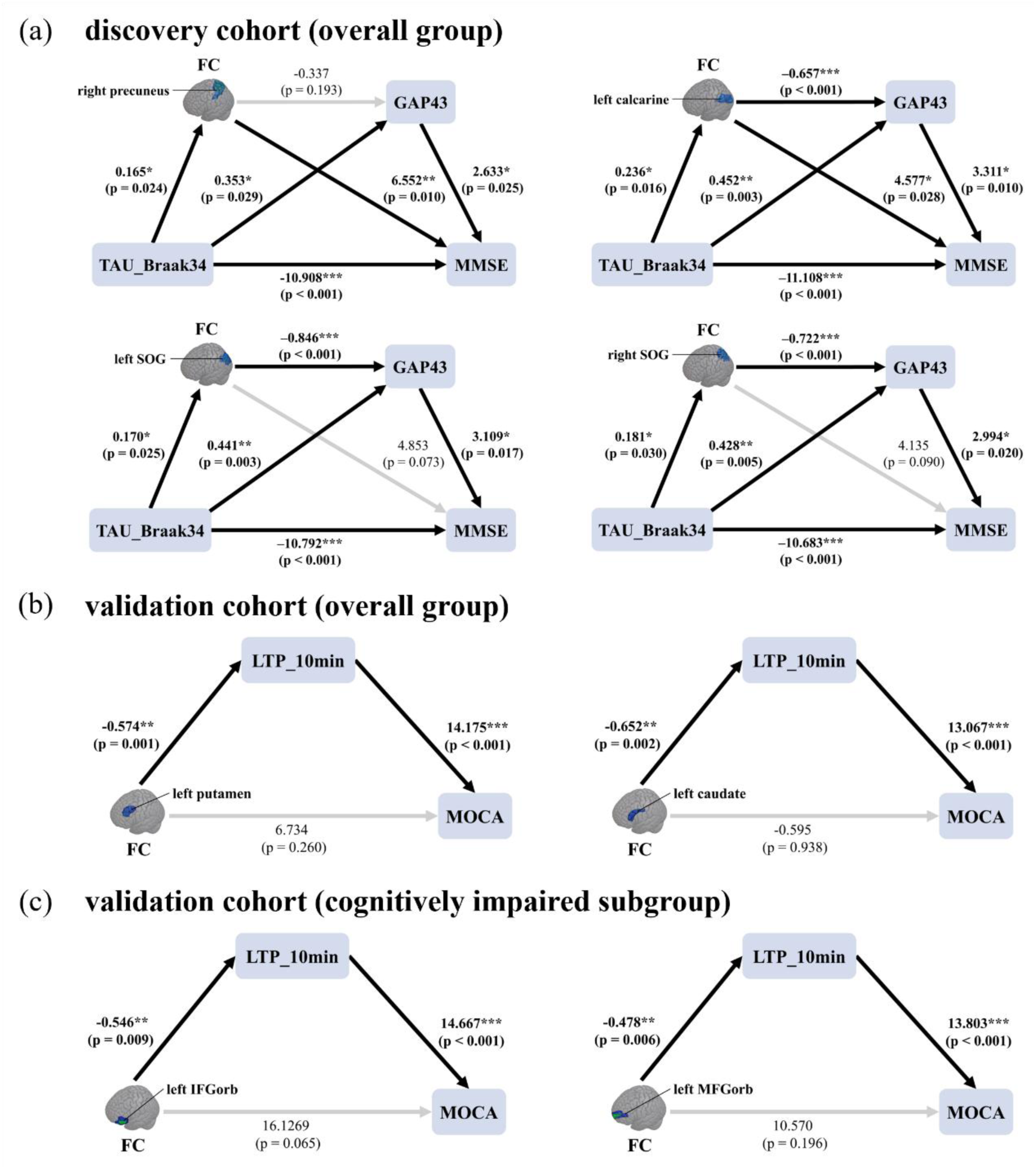
Mediation models linking functional connectivity, synaptic biomarkers, and cognitive performance. (a) Serial mediation analyses in the discovery cohort. Models illustrate the indirect association between tau pathology in Braak 3/4 regions and global cognitive performance, as assessed by the Mini-Mental State Examination (MMSE), through sequential alterations in high-NFT medial temporal lobe (MTL)-based functional connectivity (FC) and cerebrospinal fluid (CSF) GAP-43 levels. Significant serial indirect effects were identified for FC involving the calcarine cortex, precuneus, and superior occipital gyrus (SOG). (b) Simple mediation analyses in the validation cohort. Left: In the overall sample, long-term potentiation (LTP)-like plasticity measured at 10 min after stimulation (LTP-10min) significantly mediated the associations between FC and Montreal Cognitive Assessment (MoCA) scores in the left caudate nucleus (95% CI: −17.70 to −3.90) and right putamen (95% CI: −16.29 to −3.26). Right: Among participants with subjective cognitive decline (SCD) or mild cognitive impairment (MCI), LTP-10min significantly mediated the associations between FC and MoCA scores in the orbital part of the left inferior frontal gyrus (IFGorb; 95% CI: −17.44 to −2.45) and medial orbital part of the right superior frontal gyrus (95% CI: −15.70 to −2.01). Values adjacent to the arrows represent regression coefficients and *p*-values. Black arrows indicate statistically significant paths, whereas gray arrows indicate nonsignificant paths. * *p* < 0.05, ** *p* < 0.01, *** *p* < 0.001. FC, functional connectivity; GAP-43, growth-associated protein 43; MMSE, Mini-Mental State Examination; MoCA, Montreal Cognitive Assessment; SOG, superior occipital gyrus; IFGorb, orbital part of the inferior frontal gyrus; L, left; R, right.

In the validation cohort, although the full serial mediation model did not reach statistical significance, simple mediation analyses revealed significant indirect associations between FC and cognitive performance through early-phase LTP (LTP 10 min) across both in the overall sample and in the cognitively impaired subgroup (Fig. 4b).

### 3.5 Longitudinal relationship among FC, synaptic biomarkers, and cognition

CLPM analyses revealed longitudinal associations consistent with a sequential pathway linking tau pathology, FC alterations, synaptic dysfunction, and cognitive decline (Fig. 5a). First, greater baseline tau burden in Braak 1 regions predicted lower follow-up high-NFT MTL-based FC in the precuneus (*β* = −0.321, *p* = 0.022), left ITG (*β* = −0.571, *p* < 0.001, FDR-corrected *p* = 0.003) and right ITG (*β* = −0.335, *p* = 0.018). Higher baseline tau burden in Braak 3/4 and 5/6 regions also predicted lower follow-up FC in the left ITG (*β* = −0.471, *p* = 0.001 and *β* = −0.292, *p* = 0.038, respectively). Second, baseline high-NFT MTL-based FC in the right triangular part of the inferior frontal gyrus (IFGtri; *β* = −0.329, *p* < 0.001, FDR-corrected *p* = 0.013) and left precuneus (*β* = −0.183, *p* = 0.046) negatively predicted subsequent CSF GAP-43 levels. Third, higher baseline CSF GAP-43 levels predicted lower follow-up MoCA scores (*β* = −0.145, *p* = 0.001). Conversely, higher baseline MoCA scores significantly predicted greater follow-up FC in both the identified precuneus cluster (*β* = 0.231, *p* = 0.003) and the left precuneus (*β* = 0.250, *p* = 0.002).

**Fig. 5.**
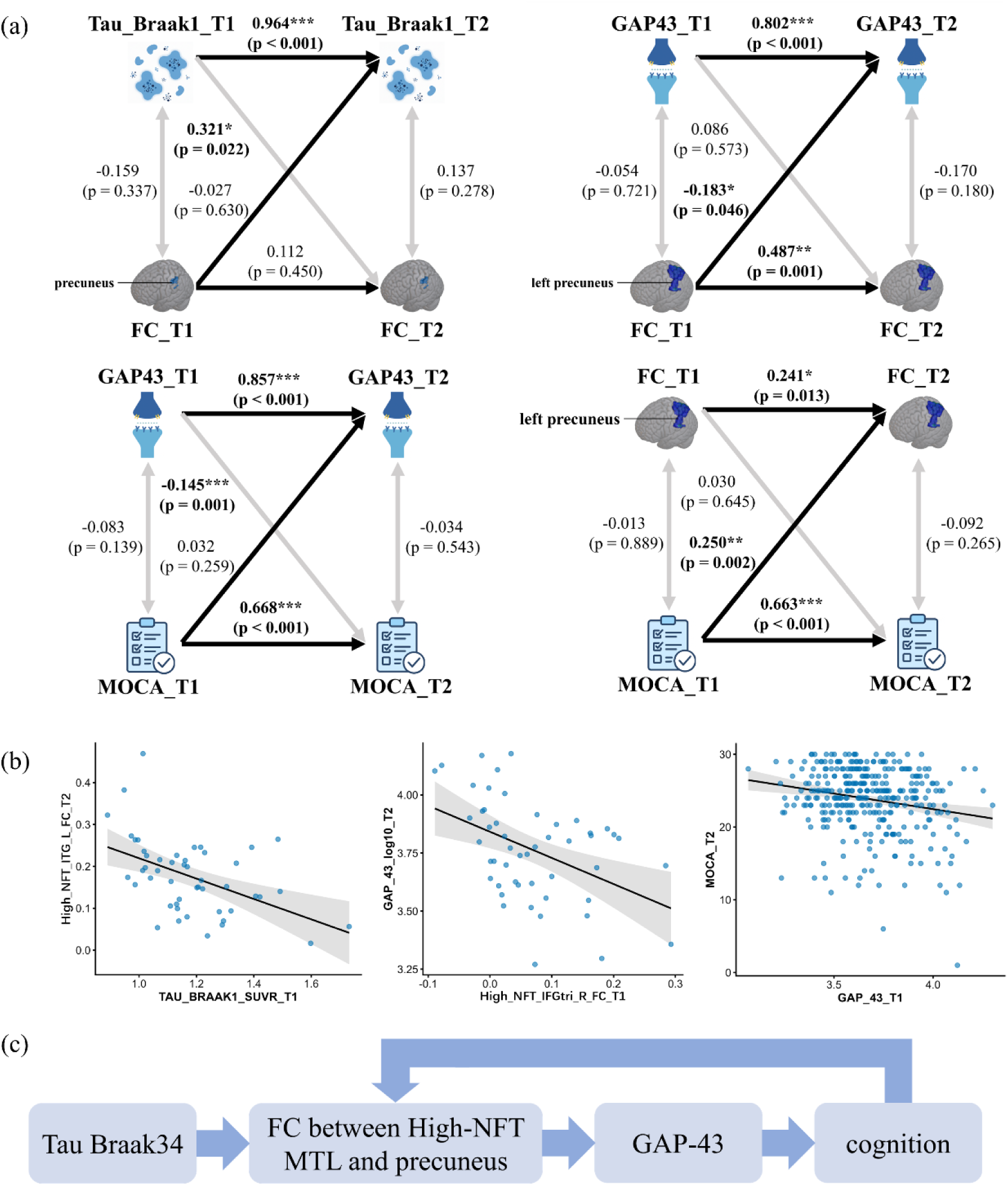
Longitudinal associations and potential functional–synaptic feedback mechanisms in the AD pathological cascade. (a) Cross-lagged panel models (CLPMs) illustrating temporal associations among tau pathology, functional connectivity (FC), synaptic biomarkers, and cognitive performance. Greater baseline tau burden in Braak 1 regions significantly predicted lower follow-up high-NFT medial temporal lobe (MTL)–precuneus FC. Baseline high-NFT MTL–left precuneus FC negatively predicted follow-up cerebrospinal fluid (CSF) GAP-43 levels. Conversely, higher baseline Montreal Cognitive Assessment (MoCA) scores positively predicted subsequent precuneus FC, whereas greater baseline CSF GAP-43 levels negatively predicted follow-up MoCA scores. (b) Scatter plots with fitted linear regression lines illustrating longitudinal associations between baseline and follow-up measures. Baseline tau burden in Braak 1 regions predicted follow-up high-NFT MTL–left inferior temporal gyrus (ITG) FC, baseline high-NFT MTL–right inferior frontal gyrus pars triangularis (IFGtri) FC predicted follow-up CSF GAP-43 levels, and baseline CSF GAP-43 levels predicted subsequent MoCA scores. (c) Schematic summary of the proposed pathological pathways. The panel illustrates a sequential pathway linking tau accumulation to cognitive decline through high-NFT MTL–precuneus FC alterations and subsequent synaptic dysfunction, and potential bidirectional relationships among network dysfunction, synaptic impairment, and cognitive decline, suggesting a possible feedback mechanism during AD progression. * *p* < 0.05, ** *p* < 0.01, *** *p* < 0.001. MoCA, Montreal Cognitive Assessment; FC, functional connectivity; ITG, inferior temporal gyrus; IFGtri, triangular part of the inferior frontal gyrus; MTL, medial temporal lobe; GAP-43, growth-associated protein 43; L, left; R, right.

Longitudinal linear regression analyses provided further support for these temporal associations (Fig. 5b). Greater baseline tau burden in Braak 1 regions significantly predicted lower follow-up FC between the high-NFT MTL region and the left ITG (*β* = −0.264, *p* < 0.001, FDR-corrected *p* = 0.023). Greater baseline high-NFT MTL-based FC in the right IFGtri was significantly associated with lower follow-up CSF GAP-43 levels (β = −0.805, *p* < 0.001, FDR-corrected *p* = 0.029). Furthermore, higher baseline CSF GAP-43 levels significantly predicted lower follow-up MoCA scores (*β* = −3.114, *p* < 0.001, FDR-corrected *p* < 0.001).

Together, these longitudinal findings suggest potential bidirectional associations among high-NFT MTL–precuneus FC, synaptic dysfunction, and cognitive performance, alongside a sequential pathway linking tau accumulation to subsequent FC alterations, synaptic dysfunction, and cognitive decline (Fig. 5c).

### 3.6 rTMS-induced changes in FC, synapse, and cognition

Following HF-rTMS targeting the L-DLPFC (Fig. 6a), paired t-tests revealed significant improvements in MoCA scores (t = 2.94, *p* = 0.007) and Auditory Verbal Learning Test–Huashan version immediate recall scores (AVLT-H-IR-S; t = 2.45, *p* = 0.022) in the overall intervention cohort. Within the SCD subgroup, AVLT-H-IR-S scores increased significantly (t = 2.71, *p* = 0.014), whereas MoCA scores showed a trend toward improvement (t = 1.93, *p* = 0.069; Fig. 6b). LTP-10min also increased significantly in both the overall cohort (t = 5.66, *p* < 0.001) and the SCD subgroup (t = 5.15, *p* < 0.001; Fig. 6c). In contrast, no significant pre-to post-intervention changes were observed in FC between either the high-NFT or low-NFT MTL seeds, and the ITG or precuneus in either group (Fig. 6d). Nevertheless, mediation analyses revealed significant indirect associations between changes in FC and changes in AVLT-H recognition scores (ΔAVLT-H-REC-S) through changes in LTP-10min (ΔLTP-10min; Fig. 6e). Specifically, significant indirect effects were observed for FC between the high-NFT MTL and right precuneus (95% CI: 0.94 to 51.21) and between the low-NFT MTL and right precuneus (95% CI: 4.39 to 78.89).

**Fig. 6.**
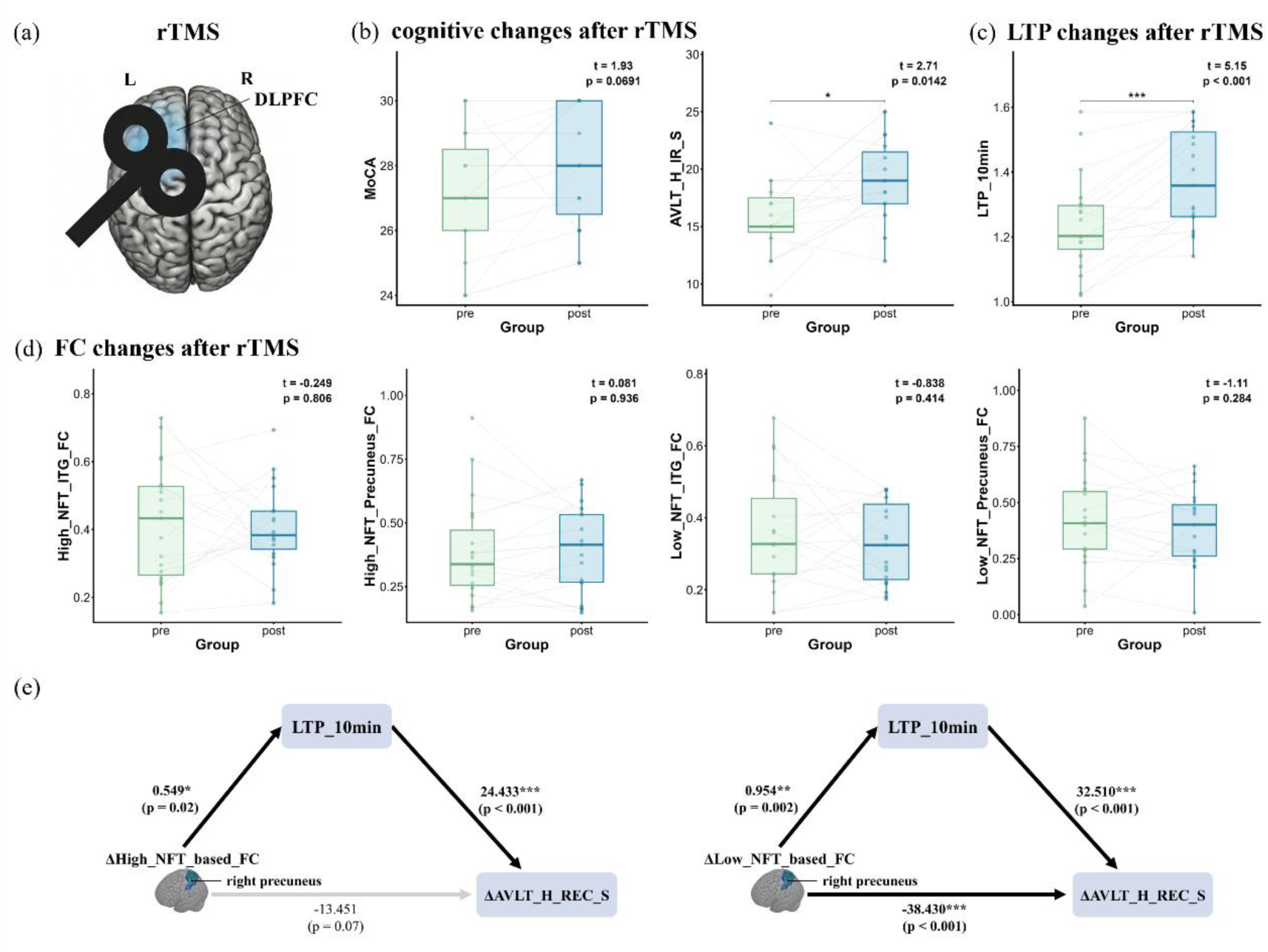
Intervention-related changes and mediation pathways in the SCD subgroup. (a) Schematic illustration of the high-frequency repetitive transcranial magnetic stimulation (HF-rTMS) protocol targeting the left dorsolateral prefrontal cortex (L-DLPFC). (b) Paired comparisons of cognitive performance before and after the intervention in the SCD subgroup. (c) Paired comparisons of LTP-10min before and after the intervention in the SCD subgroup. (d) Paired comparisons of functional connectivity (FC) between the high-NFT and low-NFT medial temporal lobe (MTL) seeds and target cortical regions, including the inferior temporal gyrus (ITG) and precuneus. (e) Mediation models in the SCD subgroup showing that changes in LTP-10min (ΔLTP-10min) mediate the association between changes in FC (ΔFC) of the high-NFT or low-NFT MTL–right precuneus connections and changes in AVLT-H recognition scores (ΔAVLT-H-REC-S). Values adjacent to the arrows represent standardized regression coefficients and corresponding P values. Black arrows indicate significant pathways, whereas gray arrows indicate nonsignificant pathways. * *p* < 0.05, ** *p* < 0.01, *** *p* < 0.001. HF-rTMS, high-frequency repetitive transcranial magnetic stimulation; L-DLPFC, left dorsolateral prefrontal cortex; MoCA, Montreal Cognitive Assessment; AVLT-H, Auditory Verbal Learning Test–Huashan version; IR-S, immediate recall score; REC-S, recognition score; LTP, long-term potentiation; FC, functional connectivity; ITG, inferior temporal gyrus; MTL, medial temporal lobe; SCD, subjective cognitive decline.

### 3.7 Association between post-mortem neuropathological Braak stages and ante-mortem FC

For FC between the high-NFT MTL region and the ITG, correlation analysis revealed a nonsignificant negative association with neuropathological Braak stage (*r* = −0.279, *p* = 0.247). Consistently, group-level comparisons showed a decreasing trend in high-NFT MTL–ITG FC with advancing Braak stage, although the differences were not statistically significant (*p* = 0.238; Supplementary Fig. 5a). In contrast, high-NFT MTL–precuneus FC exhibited a potential inverted U-shaped relationship with Braak stage. The SLM test suggested an initial increase in FC, with an estimated turning point at Braak stage 3, followed by a decline at more advanced stages. However, this nonlinear pattern was not statistically significant (SLM test, *p* = 0.228). Similarly, group-level ANCOVA showed numerically higher precuneus FC at intermediate Braak stages (3-4) than at early (1-2) and late (5-6) stages, but the group differences did not reach statistical significance (*p* = 0.159; Supplementary Fig. 5b).

## 4. DISCUSSION

The present study provides a comprehensive spatiotemporal framework for characterizing the pathological cascade of AD, linking regional tau pathology to large-scale network dysfunction, synaptic alterations, and cognitive decline. Our findings identify tau-associated FC abnormalities involving the MTL, which are linked to subsequent synaptic dysfunction and cognitive deterioration. These relationships exhibit stage-dependent patterns of FC–cognition associations and potentially bidirectional longitudinal interactions between FC and cognitive performance. Furthermore, rTMS-related cognitive improvements were associated with enhanced synaptic plasticity and altered MTL–precuneus connectivity. Taken together, these findings support a potential biological framework in which tau-associated network dysfunction and synaptic alterations contribute to cognitive decline, while suggesting reciprocal interactions between functional network integrity and cognition across the AD continuum.

### 4.1 Altered FC of high-NFT MTL regions in early AD

Our study revealed significant reductions in FC within the high-NFT MTL network, but not the low-NFT MTL network, across the AD continuum. This pattern highlights the selective vulnerability of MTL subregions and suggests that functional network disruption is closely associated with regions exhibiting substantial tau accumulation [9, 44]. The most prominent FC reductions in the high-NFT MTL network involved the ITG and precuneus. First, the ITG, a key region involved in ventral visual processing and semantic memory, is particularly vulnerable to the spread of AD-related pathology from the MTL because of its extensive anatomical connections with the entorhinal cortex [45, 46]. Synaptic loss has been documented in the ITG as early as the MCI stage, indicating its early vulnerability to AD-related neurodegeneration [47]. Elevated levels of astrocyte-derived neurotrophic factor have also been reported in the ITG during preclinical stages of AD [48, 49]. Furthermore, altered FC between the MTL and ITG has been observed in individuals with early MCI [50]. Second, the precuneus, a central hub of the DMN, plays an important role in internally directed cognitive processes, particularly episodic memory retrieval and self-referential processing [51, 52]. Its extensive functional connections with medial temporal structures support the integration of memory-related information across distributed cortical networks [53]. Previous studies have reported reduced FC involving the precuneus in individuals with AD [54]. Notably, alterations in hippocampal–precuneus FC have been observed even in the absence of substantial structural atrophy, suggesting that functional abnormalities may emerge before overt neurodegeneration [55]. Similar connectivity alterations have also been detected in individuals at increased risk of developing AD [56]. Collectively, these findings identify the ITG and precuneus as vulnerable network hubs in which functional connectivity abnormalities may precede structural neurodegeneration. Consistent with this interpretation, our longitudinal CLPM analysis demonstrated that higher baseline tau burden significantly predicted subsequent reductions in FC between high-NFT MTL regions and both the ITG and precuneus. These findings further support the utility of NFT-informed MTL subdivisions for characterizing early functional network alterations associated with AD pathology.

### 4.2 Region- and Stage-dependent associations of FC with synaptic plasticity, pathology, and clinical performance

Recent studies have highlighted the role of macroscale brain connectivity in shaping the spatial distribution of microscale synaptic degeneration [57]. Consistent with this framework, our findings across both cohorts demonstrated significant associations between high-NFT MTL-based FC and synaptic biomarkers. Specifically, the positive association between MTL–precentral gyrus FC and GAP-43 levels (which reflects molecular presynaptic remodeling [20]) may reflect a compensatory response to early AD-related pathological changes. The precentral gyrus, a key region involved in voluntary motor control, has been reported to exhibit increased coactivation during early AD, potentially reflecting neural compensation for declining functional efficiency [9, 44]. Accordingly, the association between elevated GAP-43 levels and stronger MTL–precentral gyrus FC may indicate increased synaptic remodeling associated with compensatory network reorganization [58]. In contrast, the negative association between MTL–cerebellar FC and GAP-43 levels may reflect a different pattern of network vulnerability. The cerebellum may exhibit relatively preserved local metabolic activity despite reduced integration with distributed cortical networks [59, 60]. In this context, reduced MTL–cerebellar connectivity associated with elevated GAP-43 levels may indicate impaired network integration accompanying synaptic alterations. Together, these findings suggest that AD progression is associated with region-dependent reorganization of FC, potentially involving compensatory connectivity patterns in regions such as the precentral gyrus and functional decoupling involving the cerebellum.

Linear regression analyses in both the discovery and validation cohorts revealed a stage-dependent reversal in the association between high-NFT MTL–precuneus FC and clinical performance. Stronger connectivity was associated with better cognitive performance in CN individuals but significantly worse clinical outcomes in those with advanced AD. Similar reversals were observed in the associations of both Aβ and tau burdens with MTL–precuneus FC across the two cohorts. During early AD, FC alterations emerge in association with amyloid deposition before detectable structural or metabolic abnormalities [56]. As pathological burden increases, progressive AD-related changes disrupt the normal relationship between FC and cognitive performance [48, 49]. Further evidence of stage-dependent connectivity patterns emerged from correlation analyses in the validation cohort. Despite an overall negative association in synaptic efficacy, early-phase LTP-like plasticity (which captures neurophysiological synaptic efficacy [24]) remained positively associated with FC involving the left angular gyrus and left posterior cingulate cortex in individuals with MCI. These findings suggest that increased FC during early disease stages may reflect adaptive or compensatory processes that help maintain cognitive function, whereas increased connectivity at more advanced stages indicate maladaptive network activity [61]. This interpretation is further supported by the inverted U-shaped trajectory of high-NFT MTL–precuneus FC across postmortem Braak stages, consistent with a cascading network failure model in which early pathological changes are accompanied by transient hyperconnectivity, followed by progressive network dysfunction and eventual connectivity decline [62]. These findings highlight dynamic, stage-dependent changes in brain–behavior relationships rather than a fixed association between FC and cognitive performance. The observed reversal may reflect a progressive disruption of normal brain–behavior coupling, potentially marking a transition from compensatory network activity to functional impairment [63]. However, further validation in larger AD cohorts is warranted, particularly given the limited sample size of the AD subgroup.

In our longitudinal CLPM and regression analyses, the precuneus and IFGtri emerged as key regions in which FC predicted subsequent changes in synaptic biomarkers. The predictive association involving the precuneus further highlights its potential role as a critical hub linking functional network disruption to synaptic dysfunction. The IFGtri, an important component of frontal control and salience-related networks, contributes to higher-order cognitive and affective processing and facilitate the integration of external information with internal states [64, 65]. Previous studies have reported altered activation and enhanced responses to familiar stimuli in this region in individuals with AD, suggesting compensatory recruitment of frontal regions [66]. Our findings further suggest that disrupted functional coupling between the MTL and IFGtri may contribute to synaptic instability, consistent with evidence linking inferior frontal degeneration to increased clinical vulnerability [67]. Studies in AD mouse models have shown that baseline FC predicts alterations in activity-dependent markers associated with synaptic dysfunction and loss [68]. Human studies have also demonstrated that synaptic alterations are associated with the underlying FC architecture, with more pronounced synaptic changes occurring in regions more strongly connected to tau epicenters [27, 57]. Meanwhile, synaptic systems may undergo compensatory plasticity and dynamic remodeling in response to functional disruption and network stress [69]. Collectively, these findings suggest that FC abnormalities may precede widespread synaptic deterioration.

### 4.3 A functional-to-synaptic pathological cascade linking tau accumulation to cognitive decline

Significant serial mediation effects linking tau pathology to cognitive performance through sequential alterations in FC and synaptic biomarkers were identified not only in the precuneus but also in occipital regions, including the calcarine cortex and SOG. These findings highlight the involvement of posterior cortical regions in linking pathological burden to cognitive decline. The calcarine cortex is affected across multiple stages of AD and exhibits structural alterations associated with molecular pathways implicated in neurodegeneration [70, 71]. Similarly, structural alterations in the SOG have been associated with disease progression and the risk of conversion from MCI to AD [72]. These observations suggest that posterior visual regions may contribute to broader network-level processes underlying AD progression.

The identified serial mediation pathway provides a potential multilevel framework linking tau accumulation to cognitive impairment through functional network disruption and subsequent synaptic alterations, which further supported the findings from the longitudinal CLPM and regression analyses. Previous studies have demonstrated that pathological tau propagates along functionally interconnected brain networks, progressively disrupting connectivity within these pathways [14, 15, 73]. Additionally, elevated levels of the presynaptic protein GAP-43 have been associated with accelerated tau accumulation and propagation across functionally connected brain regions [74]. As synaptic dysfunction is a central feature of AD, these alterations may contribute substantially to cognitive impairment [75]. Furthermore, recent research has highlighted stage-dependent relationships between protein pathology and synaptic-functional integrity, demonstrating associations of presynaptic biomarkers, including GAP-43, with amyloid-β burden, FC alterations, and early cognitive decline [76]. Taken together, our findings provide a spatiotemporal framework connecting regional tau accumulation with progressive high-NFT MTL-based functional network disruption, synaptic dysfunction, and cognitive deterioration across the AD continuum.

Furthermore, this functional-to-synaptic pathway may also contribute to the neural mechanisms underlying HF-rTMS-related cognitive improvement. Our mediation analysis revealed that enhanced synaptic plasticity significantly mediated the association between precuneus-related FC and cognitive gains following HF-rTMS, despite the absence of significant overall changes in FC. Whereas previous studies have largely examined FC and synaptic excitability independently, our findings provide preliminary interventional evidence supporting a potential role for synaptic plasticity in linking functional network modulation to cognitive improvement [74]. These results provide further support for the proposed functional-to-synaptic cascade and highlight high-NFT MTL–precuneus circuits as promising targets for early neuromodulatory interventions.

In addition, our CLPM analysis suggests a potential feedback loop involving FC, synaptic alterations, and cognitive decline. Specifically, baseline FC between the high-NFT MTL and left precuneus predicted subsequent GAP-43 levels, which were associated with later cognitive decline. In turn, cognitive impairment may contribute to further deterioration of functional network integrity. Previous studies have demonstrated that FC involving precuneus-related networks predicts individual differences in memory and global cognitive performance [77, 78]. Extending these findings, our analysis identified a reverse longitudinal association, whereby higher baseline MoCA scores positively predicted subsequent FC between the high-NFT MTL and left precuneus, suggesting a bidirectional relationship between cognitive performance and functional network integrity. One possible interpretation involves activity-dependent plasticity, through which the sustained cognitive engagement may help maintain functional integration across distributed brain networks, and vice versa [79–81]. Notably, this reverse association was specific to FC involving high-NFT MTL subfields, suggesting that these tau-vulnerable regions may be particularly sensitive to cognitive status. This pattern is consistent with the cascading network failure model, which proposes that AD progression involves mutually reinforcing interactions among pathological and functional alterations within distributed brain networks [82]. Within this framework, the left precuneus may represent a critical network hub where early functional disturbances contribute to progressive network dysfunction. In summary, our findings suggest that interactions between the high-NFT MTL and DMN hubs may be dynamically associated with cognitive performance, potentially contributing to a self-reinforcing cycle of functional network deterioration over time.

### 4.4 Limitations

Several limitations of this study should be acknowledged. First, the diagnostic compositions differed between the two cohorts. The discovery cohort included CN individuals, patients with MCI, and patients with AD dementia, whereas the validation cohort comprised CN individuals and participants with SCD or MCI, without an AD dementia group. These differences in disease-stage coverage may limit direct cross-cohort comparisons across the full AD continuum. Second, different synaptic biomarkers were used in the two cohorts. Although CSF GAP-43 and LTP-like plasticity both provide information about synaptic function, they reflect distinct biological processes. These differences may contribute to inconsistencies between cohorts and limit the direct comparability of findings. Third, postmortem neuropathological validation was conducted in a relatively small, separate subset of participants, potentially limiting the generalizability of these findings. Future studies integrating antemortem neuroimaging with subsequent neuropathological assessments in the same individuals would provide more direct validation of the proposed relationships. Last but not least, the potential functional–synaptic feedback mechanism was inferred from two-wave longitudinal data, which are insufficient to establish reciprocal causal relationships or fully characterize dynamic feedback processes. Future studies incorporating multiple longitudinal assessments are needed to clarify the temporal dynamics and potential bidirectional interactions among FC, synaptic dysfunction, and cognitive decline.

## 5. CONCLUSIONS

This study provides evidence for a potential pathological cascade in AD using an NFT-informed MTL framework, suggesting that tau accumulation may precede FC alterations involving the ITG and precuneus. In addition, mediation and longitudinal analyses support a sequential pathway linking tau pathology to cognitive decline through high-NFT MTL-based network dysfunction and subsequent synaptic alterations, alongside potential bidirectional relationships between FC and cognitive performance involving the precuneus. Furthermore, rTMS-induced cognitive improvements were associated with changes in MTL–precuneus FC through enhanced synaptic plasticity. Collectively, these findings highlight high-NFT MTL-based FC as a promising candidate biomarker for early AD-related network dysfunction and identify tau-vulnerable MTL circuits as potential targets for therapeutic intervention.

## Data Availability

Data used in the discovery cohort were obtained from the Alzheimer's Disease Neuroimaging Initiative (ADNI) database (https://adni.loni.usc.edu/) and are openly available to qualified researchers upon application and registration. The validation cohort and rTMS intervention data generated from Jiangsu Shengze Hospital are available upon reasonable request to the authors.

## Acknowledgments

This work was supported by Central Guidance Fund for Local Scientific and Technological Development (ZYYD2026QY10), Special Fund for Talents of Children’s Hospital Affiliated to Zhejiang University School of Medicine (Y024016), and National Natural Science Foundation of China (No. 82372582, No. 82603967, No. 82671848).

## Author Contribution

Z.Y.Z., and Y.S. conceptualized and designed the study. Y.J.Z. wrote the original draft of the manuscript. All authors contributed to the review and editing, and they approved the final version of the manuscript. Z.Y.Z, Y.S., and F.H. acquired funding. Y.H.M and Q.L performed the experimental investigations. Y.J.Z and K.Y.L conducted the formal analysis and contributed to data visualization. Z.Y.Z and D.W. supervised the project.

