## Supplementary Material for "Tau-vulnerable medial temporal lobe connectivity drives a functional-to-synaptic cascade in Alzheimer’s disease"

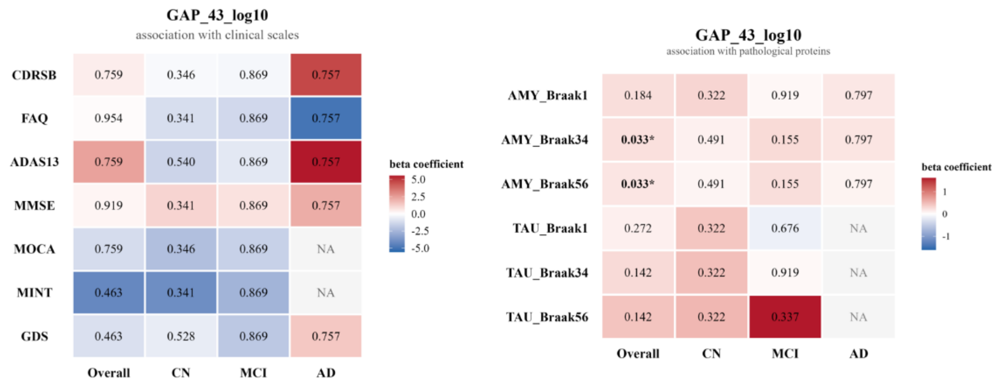


**Supplementary Fig. 1. Associations of CSF GAP-43 levels with clinical performance and pathological burden.** (a) Heatmap illustrating associations between log10-transformed cerebrospinal fluid (CSF) GAP-43 levels and clinical assessment scores in the overall cohort and diagnostic subgroups (CN, MCI, and AD). Although no associations survived false discovery rate (FDR) correction, stage-dependent reversals in the direction of associations were observed for several clinical measures, including CDR-SB and ADAS13. (b) Heatmap illustrating associations between CSF GAP-43 levels and amyloid-β (Aβ) and tau pathological burden across Braak-stage-defined regions. In the overall cohort, CSF GAP-43 levels were significantly positively associated with Aβ burden in Braak III/IV ($\beta = 0.24$, FDR-corrected $p = 0.033$) and V/VI ($\beta = 0.26$, FDR-corrected $p = 0.033$) regions. No significant associations were observed with tau pathology or within individual diagnostic subgroups. Color bars represent standardized regression coefficients (β), and values within cells indicate FDR-corrected p-values. *$p < 0.05$. CDR-SB, Clinical Dementia Rating Sum of Boxes; FAQ, Functional Activities Questionnaire; ADAS13, 13-item Alzheimer's Disease Assessment Scale–Cognitive Subscale; MMSE, Mini-Mental State Examination; MoCA, Montreal Cognitive Assessment; MINT, Multilingual Naming Test; GDS, Geriatric Depression Scale; AMY, amyloid-β; CN, cognitively normal; MCI, mild cognitive impairment; AD, Alzheimer's disease; FDR, false discovery rate.

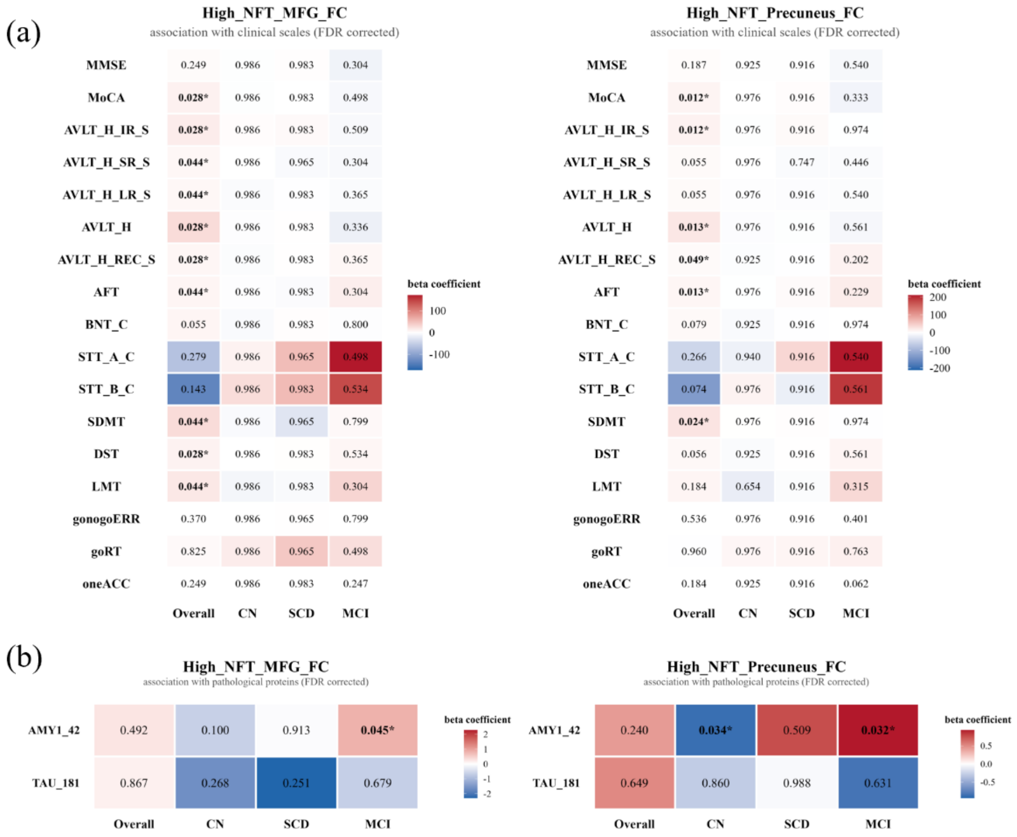


**Supplementary Fig. 2. Associations between functional connectivity, cognitive performance, and pathological biomarkers in the validation cohort.** (a) Associations of functional connectivity (FC) between the high-NFT medial temporal lobe (MTL) and the middle frontal gyrus (MFG) or precuneus with cognitive performance across diagnostic groups. (b) Associations of amyloid-β1–42 (Aβ1–42) and phosphorylated tau at threonine 181 (p-tau181) concentrations with FC measures. Serum biomarker concentrations were scaled to increments of 1,000 pg/L, and the regression coefficients (β) represent changes in the dependent variable associated with each 1,000 pg/L increase in biomarker concentration. Values within cells indicate false discovery rate (FDR)-corrected p values, whereas colors represent regression coefficients (β). Asterisks denote statistically significant associations (* $p < 0.05$, FDR-corrected). MFG, middle frontal gyrus; FC, functional connectivity; AVLT-H, Auditory Verbal Learning Test–Huashan version; IR-S, immediate recall score; SR-S, short-delay recall score; LR-S, long-delay recall score; REC-S, recognition score; AFT, Animal Fluency Test; BNT-C, Boston Naming Test–Chinese version; STT-A-C/STT-B-C, Shape Trail Test A/B–Chinese version; SDMT, Symbol Digit Modalities Test; DST, Digit Span Test; LMT, Logical Memory Test; gonogoERR, Go/No-Go error rate; goRT, Go reaction time; oneACC, One-Card Learning accuracy; AMY1_42, amyloid-β1–42; TAU_181, phosphorylated tau at threonine 181.

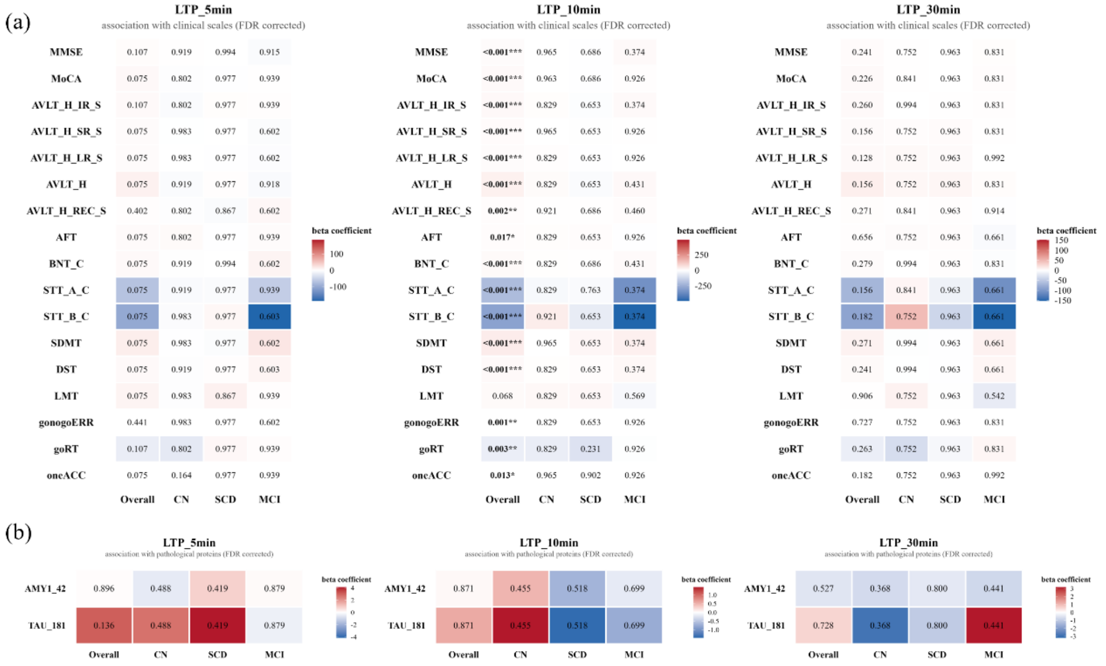


**Supplementary Fig. 3. Associations between synaptic plasticity, cognitive performance, and pathological biomarkers in the validation cohort.** (a) Associations of long-term potentiation (LTP)-like plasticity measured at 5, 10, and 30 min after stimulation (LTP-5min, LTP-10min, and LTP-30min, respectively) with cognitive performance across diagnostic groups. (b) Associations of amyloid-β1–42 (Aβ1–42) and phosphorylated tau at threonine 181 (p-tau181) concentrations with LTP-like plasticity measures. Serum biomarker concentrations were scaled to increments of 1,000 pg/L, and the regression coefficients (β) represent changes in the dependent variable associated with each 1,000 pg/L increase in biomarker concentration. Values within cells indicate false discovery rate (FDR)-corrected p values, whereas colors represent regression coefficients (β). Asterisks denote statistically significant associations: * $p < 0.05$, ** $p < 0.01$, and *** $p < 0.001$. LTP, long-term potentiation; AVLT-H, Auditory Verbal Learning Test–Huashan version; IR-S, immediate recall score; SR-S, short-delay recall score; LR-S, long-delay recall score; REC-S, recognition score; AFT, Animal Fluency Test; BNT-C, Boston Naming Test–Chinese version; STT-A-C/STT-B-C, Shape Trail Test A/B–Chinese version; SDMT, Symbol Digit Modalities Test; DST, Digit Span Test; LMT, Logical Memory Test; gonogoERR, Go/No-Go error rate; goRT, Go reaction time; oneACC, One-Card Learning accuracy; AMY1_42, amyloid-β1–42; TAU_181, phosphorylated tau at threonine 181.

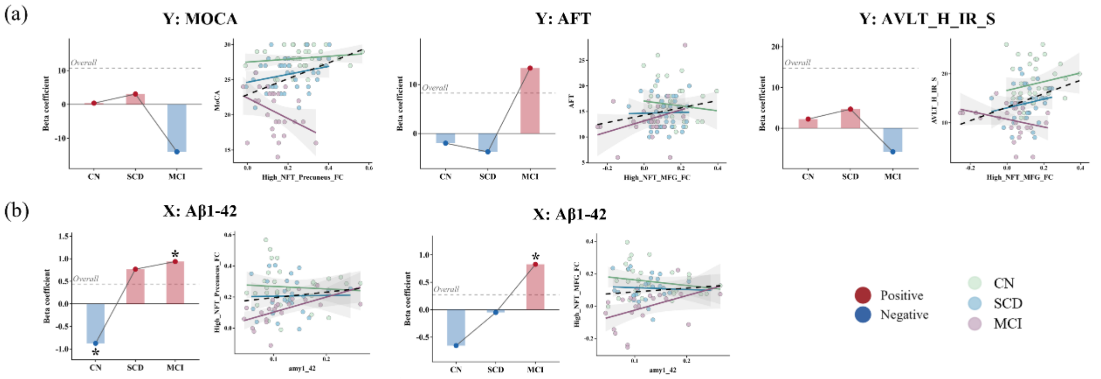


**Supplementary Fig. 4. Stage-dependent shifts in associations between high-NFT functional connectivity and clinical or pathological markers in the validation cohort.** (a) Changes in regression coefficients (*β*) for the associations between high-NFT medial temporal lobe (MTL)-based functional connectivity (FC) and cognitive performance across diagnostic groups. (b) Changes in regression coefficients (*β*) for the associations between serum amyloid-β1–42 (Aβ1–42) levels and high-NFT MTL-based FC across diagnostic groups. Serum Aβ1–42 concentrations were scaled to units of 1,000 pg/L, and regression coefficients represent changes in FC per 1,000 pg/L increase in Aβ1–42 concentration. Notably, circulating Aβ1–42 levels may not directly reflect cerebral amyloid plaque burden. Bars represent regression coefficients, and dots indicate group-specific estimates. These plots illustrate potential stage-dependent shifts in the direction and magnitude of associations across the validation cohort. Asterisks (*) indicate statistically significant associations ($p < 0.05$). MFG, middle frontal gyrus; FC, functional connectivity; LTP, long-term potentiation; MoCA, Montreal Cognitive Assessment; AVLT, Auditory Verbal Learning Test; AFT, Animal Fluency Test.

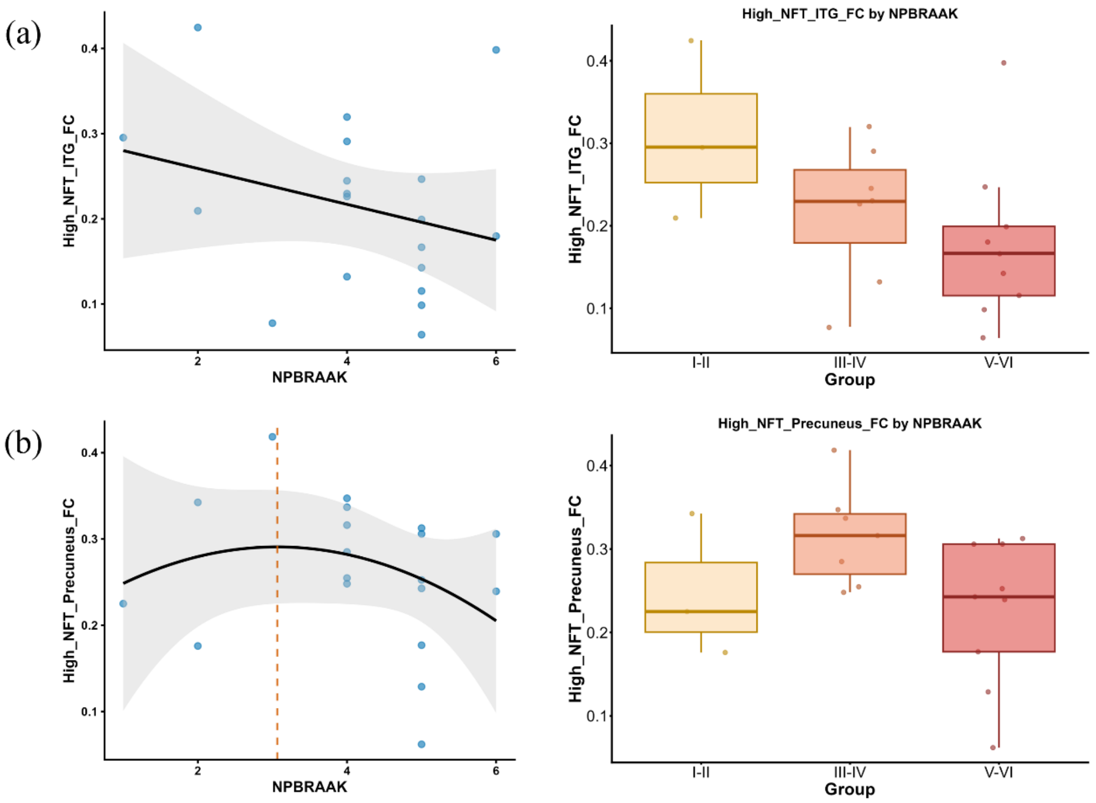


**Supplementary Fig. 5 Associations between high-NFT medial temporal lobe-based functional connectivity and neuropathological Braak stages.** (a) Associations between high-NFT medial temporal lobe (MTL)–inferior temporal gyrus (ITG) functional connectivity (FC) and neuropathological Braak stage. (b) Associations between high-NFT MTL–precuneus FC and neuropathological Braak stage. Left panels show scatter plots with fitted linear or nonlinear regression curves across Braak stages 1-6. Right panels present group-level comparisons of FC across early (Braak 1-2), intermediate (Braak 3-4), and advanced (Braak 5-6) neuropathological stages. The observed patterns were exploratory, as neither the associations nor the group differences reached statistical significance. ITG, inferior temporal gyrus; MTL, medial temporal lobe; FC, functional connectivity; NFT, neurofibrillary tangle.

**Supplementary Table 1. Demographic and clinical characteristics of participants across CN, SCD, and MCI groups in the validation cohort.**

| | CN (N=37) | SCD (N=35) | MCI (N=28) | $\chi^2$ | $p$ |
|---|---|---|---|---|---|
| Sex(F/M) | 28/9 | 25/10 | 23/5 | 0.98 | 0.612 |
| Age | 67.00 (7.00) | 64.00 (8.00) | 70.00 (5.50) | 5.88 | 0.001 |
| Education | 9.00 (3.00) | 9.00 (3.00) | 9.00 (3.00) | 0.13 | 0.938 |

| | | | | | |
|---|---|---|---|---|---|
| MMSE (N=98) | 30.00 (0.00) | 29.00 (2.00) | 26.50 (3.00) | 44.26 | <0.001 |
| MoCA (N=98) | 28.00 (2.00) | 26.00 (3.00) | 21.00 (4.75) | 59.95 | <0.001 |
